## Appendix Table 1 for "COVID-19 prevention behaviour over time in Australia: Patterns and long-term predictors from April to July 2020"

**Table 1. Descriptive characteristics of analysis sample (N=1843) and full sample invited for follow-up (N=3216) at baseline (April)**

| **Characteristic** | | **Analysis sample (N=1843) n (%)** | **Full sample (N=3216) n (%)** |
| --- | --- | --- | --- |
| Age group | |  |  |
|  | 18 to 25 years | 353 (19.2%) | 800 (24.9%) |
|  | 26 to 40 years | 528 (28.6%) | 930 (28.6%) |
|  | 41 to 55 years | 462 (25.1%) | 737 (22.9%) |
|  | 56 to 90 years | 500 (27.1%) | 749 (23.3%) |
| Gender | |  |  |
|  | Male | 487 (26.4%) | 1120 (35.0%) |
|  | Female | 1322 (71.7%) | 2033 (63.5%) |
|  | Other/prefer not to say | 34 (1.8%) | 48 (1.5%) |
| Educational attainment* | |  |  |
|  | Less than university | 496 (26.9%) | 971 (30.3%) |
|  | University | 1347 (73.1%) | 2230 (69.7%) |
| State/territory of residence | |  |  |
|  | Australian Capital Territory | 58 (3.1%) | 95 (3.0%) |
|  | Northern Territory | 7 (0.4%) | 16 (0.5%) |
|  | Victoria | 291 (15.8%) | 495 (15.5%) |
|  | New South Wales | 937 (50.8%) | 1664 (52.0%) |
|  | Queensland | 258 (14.0%) | 447 (14.0%) |
|  | Western Australia | 130 (7.1%) | 221 (6.9%) |
|  | South Australia | 84 (4.6%) | 143 (4.5%) |
|  | Tasmania | 78 (4.2%) | 120 (3.7%) |
| Residential area remoteness^ | |  |  |
|  | Major cities | 1374 (74.6%) | 2399 (75.5%) |
|  | Regional and remote | 467 (25.4%) | 780 (24.5%) |
| Socioeconomic status, mean IRSAD quintile (SD) | | 3.66 (1.40) | 3.67 (1.39) |
| Born in Australia | | 1405 (76.2%) | 2425 (75.8%) |
| English primary language | | 1774 (96.3%) | 3025 (94.5%) |
| Aboriginal or Torres Strait Islander | |  |  |
|  | Yes | 31 (1.7%) | 57 (1.8%) |
|  | No | 1796 (97.4%) | 3109 (97.1%) |
|  | Did not respond | 16 (0.9%) | 35 (1.1%) |
| Chronic health conditions | |  |  |
|  | None | 912 (49.5%) | 1683 (52.3%) |
|  | One | 537 (29.1%) | 910 (28.3%) |
|  | Two or more | 394 (21.4%) | 623 (19.4%) |
| Health literacy adequacy^†^ | | 1695 (92.0%) | 2880 (90.0%) |
| Self-Reported General Health | |  |  |
|  | Poor | 68 (3.7%) | 102 (3.2%) |
|  | Fair | 248 (13.5%) | 442 (13.8%) |
|  | Good | 629 (34.1%) | 1099 (34.3%) |
|  | Very good | 667 (36.2%) | 1144 (35.7%) |
|  | Excellent | 231 (12.5%) | 414 (12.9%) |

*Notes:* The analysis sample comprised participants from our prospective longitudinal study who provided responses to the baseline survey (April) and at least one subsequent follow-up survey (N=1,843)*.* ^Remoteness indicators are based on 2016 ABS data, and as such, individuals who reside in newer postcodes established after 2016 (n=2 in analysis sample; n=37 in full sample) are missing data on this variable. ^†^Based on Single Item Literacy Screener (SILS): How confident are you with filling out medical forms by yourself: not at all, a little bit, somewhat, quite a bit, extremely. “Not at all” response categorised as inadequate health literacy.

**Table 2. Sensitivity analyses of pairwise comparisons between the April (baseline) and subsequent surveys on distancing and hygiene component scores (i.e., ‘stay at home’ behaviour not included in the PCA). Values are presented as estimated mean differences from the fixed portion of the linear mixed models, and can be interpreted as standard deviation units.**

| Pairwise comparisons to April Survey (baseline) | Component 1: Distancing  Estimated mean difference (95% CI); p-value | Component 2: Hygiene  Estimated mean difference (95% CI); p-value |
| --- | --- | --- |
| May Survey | -0.28 (-0.34, -0.23), p<.001 | -0.14 (-0.18, -0.09), p<.001 |
| June Survey | -0.69 (-0.77, -0.61), p<.001 | -0.14 (-0.19, -0.09), p<.001 |
| July Survey | -0.80 (-0.88, -0.72), p<.001 | -0.14 (-0.20, -0.09), p<.001 |
